# Scoring consistency checks for the Clinician Administered PTSD Scale (CAPS-IV & CAPS-V)

**DOI:** 10.1101/2024.07.09.24310039

**Authors:** Jonathan Rabinowitz, Alon A. Rabinowitz, Sara Freedman

## Abstract

**Background:** The CAPS is regarded as the “gold standard” in PTSD assessment. It is a structured interview that yields a categorical diagnosis of PTSD and also a measure of the severity of PTSD symptoms. Since a PTSD diagnosis in some settings could be connected to getting benefits, scoring inconsistencies may be more abundant with this rating scale as they not only reflect raters carelessness but intentional inaccurate reporting by the subject.

**Objective:** The objective of the current effort was to derive rating consistency flags for the CAPS-IV.

**Methods:** We deconstructed CAPS scoring instructions and anchors to identify potential scoring inconsistency flags. These inconsistency flags were reviewed and confirmed by expert raters. To test the ability of the flags to identify careless responses the flags were applied to Monte Carlo simulated data of 100,000 CAPS administrations.

**Results:** Twelve flags were derived (presented in Table 1). Two flags applied to most of the 17 symptom items (Flag 1: Frequency=0 & Intensity>0 and Flag 2: Frequency>0 & Intensity=0). The remaining 10 flags pertained to individual items. Five flags were rated as “High” flags, representing very probably or definitely incorrect rating, one as medium, reflecting probably incorrect rating. Flags were raised for 95% of the Monte Carlo simulated CAPS administrations, 78% of the administrations had 4 or more flags and 60% 5 or more.

**Conclusions:** Scoring consistency flags for the CAPS may be useful in the quest to improve reliability and validity of clinical trials. Modified flags are currently being developed to cover the CAPS-V. Further testing of flags using clinical trial data is planned to examine their potential impact on signal detection.

## Introduction

The CAPS is regarded as the “gold standard” in PTSD assessment. It is a structured interview that yields a categorical diagnosis of PTSD and also a measure of the severity of PTSD symptoms. It can be administered in 30-60 minutes by a trained rater.

**Table 1.**
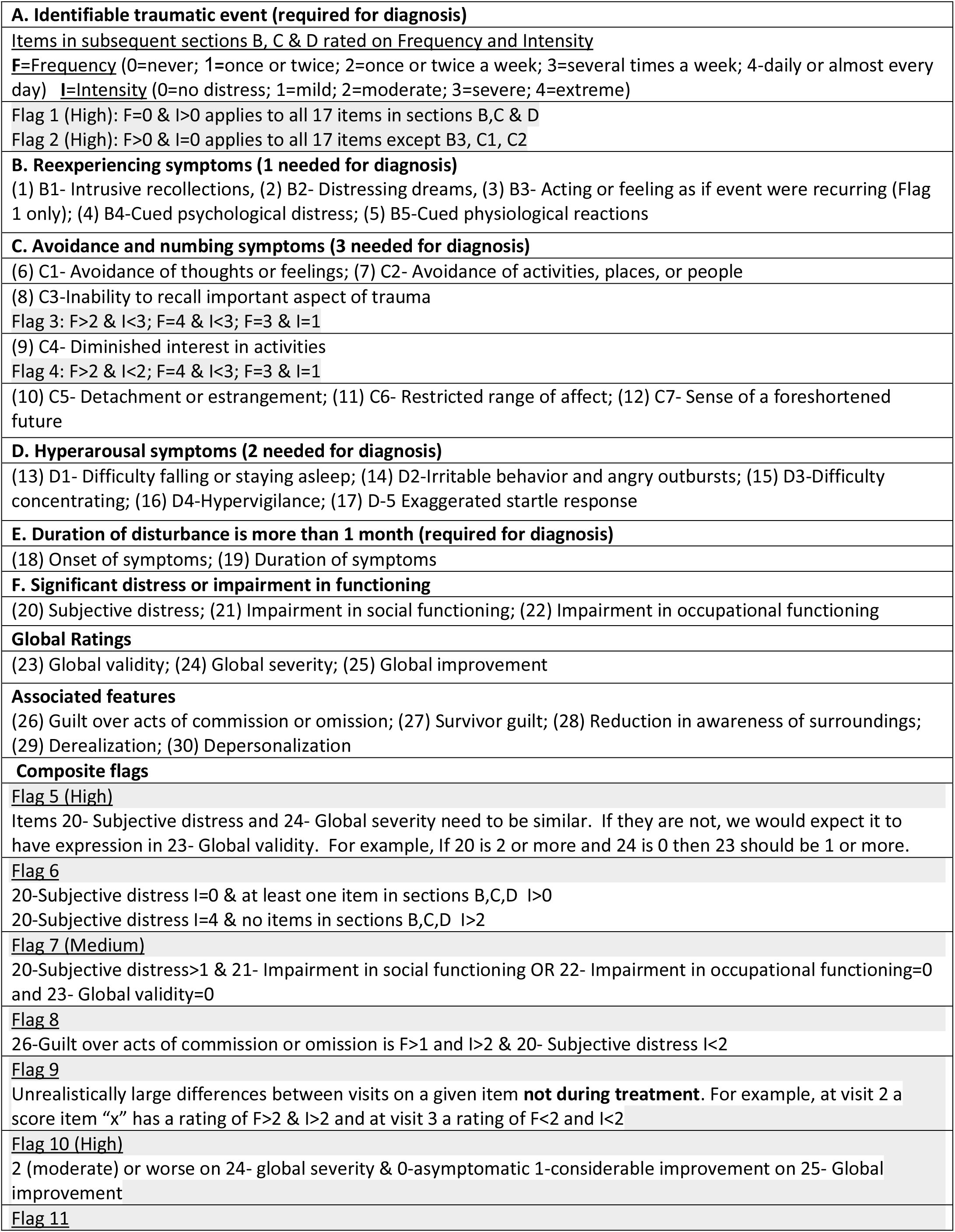
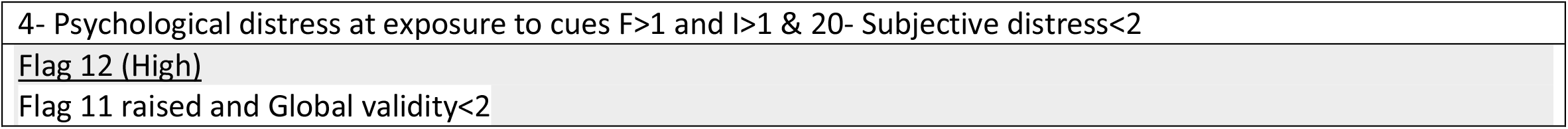
List of CAPS IV items and inconsistency FLAGS.

To improve the reliability and validity of measurement in clinical trials, we previously developed consistency checks “flags” for the Montgomery-Asberg Depression Rating Scale (MADRS)(Rabinowitz et al., 2019, 2022), Positive and Negative Syndrome Scale (PANSS) (Rabinowitz et al,, 2017; 2021), Personal and Social Performance scale (PSP) (Rabinowitz et al, 2021), the Young Mania Rating Scale (YMRS) (Rabinowitz et al, 2024).

Since a PTSD diagnosis in some settings could be connected to getting benefits, scoring inconsistencies may be more abundant with this rating scale as they not only reflect raters carelessness but intentional inaccurate reporting by the subject.

## Aim

The objective of the current effort was to derive consistency flags for the CAPS-IV.

## Methods

The first 17 items of CAPS IV elicits ratings on Frequency (0=never; 1=once or twice; 2=once or twice a week; 3=several times a week; 4-daily or almost every day) and intensity (0=no distress; 1=mild; 2=moderate; 3=severe; 4=extreme) of symptoms and are used to compute a total severity score. The next 4 items measure duration, subject distress and functional impairment. A scoring algorithm is applied to these 21 items to arrive at a diagnosis. The CAPS also includes 3 global ratings and ratings of 5 associated features.

We deconstructed CAPS scoring instructions and anchors to identify potential scoring inconsistency flags. These inconsistency flags were reviewed and confirmed by expert raters.

To test the ability of the flags to identify careless responses the flags were applied to Monte Carlo simulated data of 100,000 CAPS administrations.

## Results

Twelve flags were derived (presented in Table 1). Two flags applied to most of the 17 symptom items (Flag 1: Frequency=0 & Intensity>0 and Flag 2: Frequency>0 & Intensity=0). The remaining 10 flags pertained to individual items. Five flags were rated as “High” flags, representing very probably or definitely incorrect rating, one as medium, reflecting probably incorrect rating. Flags were raised for 95% of the Monte Carlo simulated CAPS administrations, 78% of the administrations had 4 or more flags and 60% 5 or more. Two high flags, Flag 1 and 2 were raised in more than 85% of the administrations. Table 2 presents flags for both the CAPS-IV and CAPS-V and a comparison of the items on each scale.

**Table 2.**
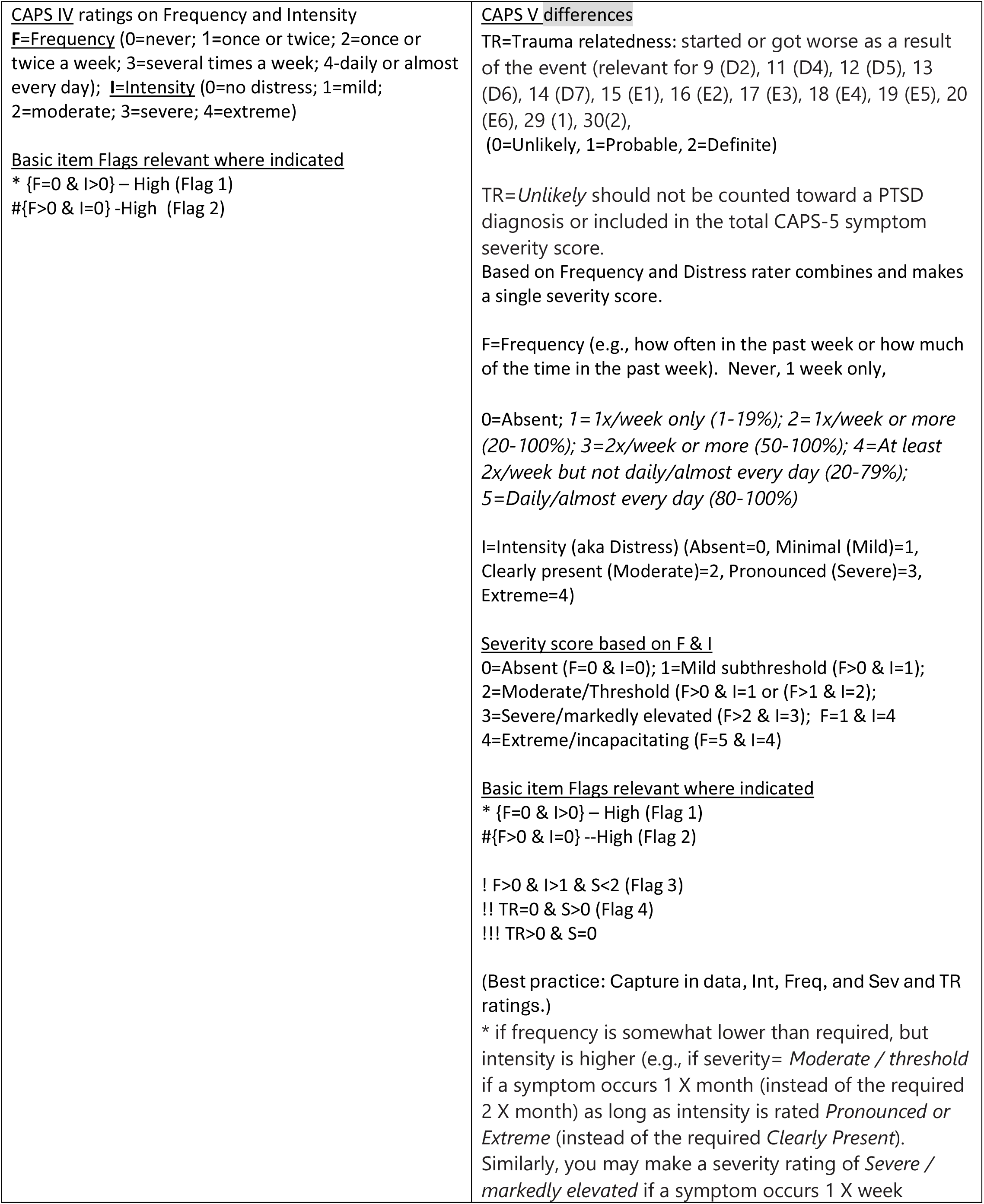
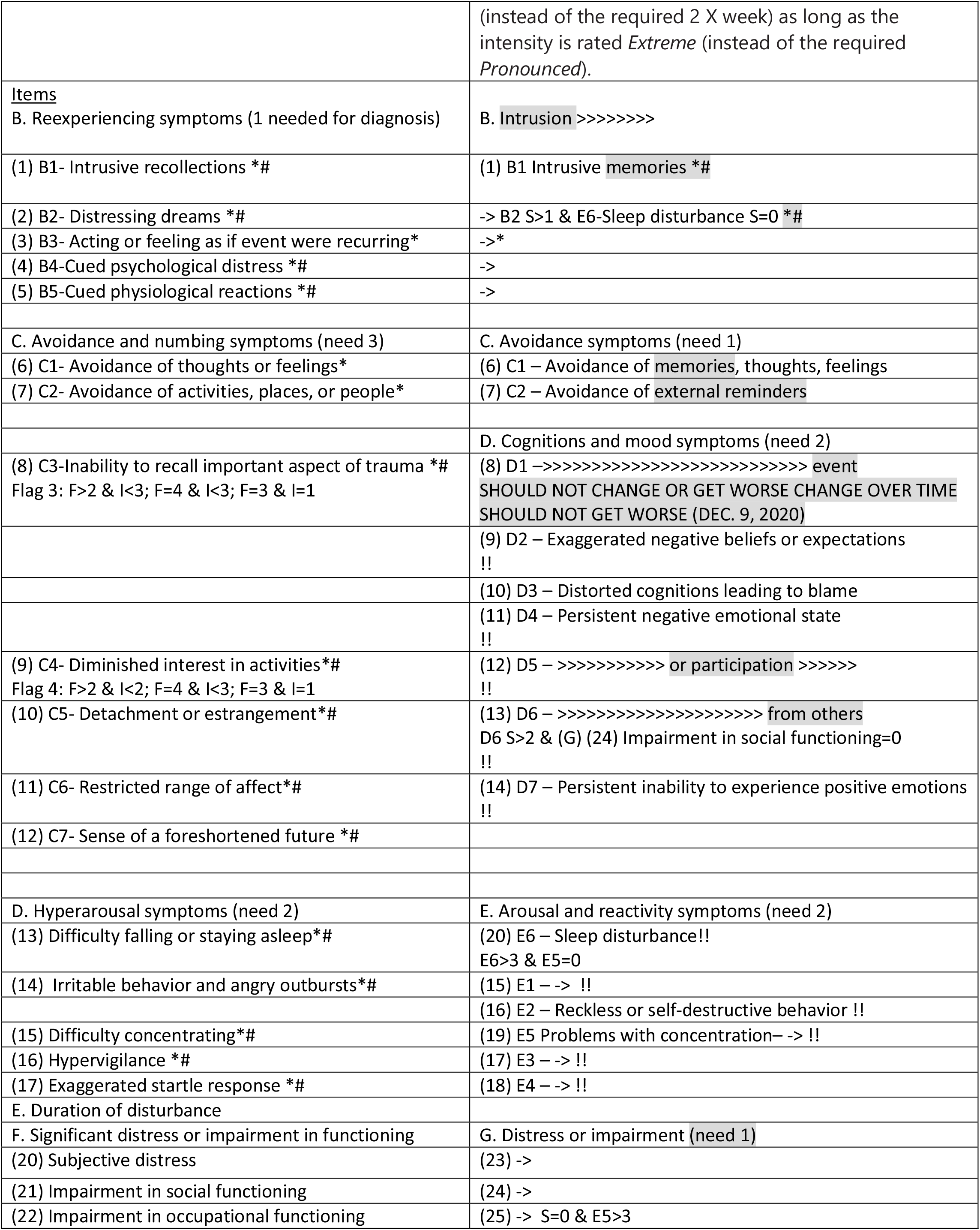
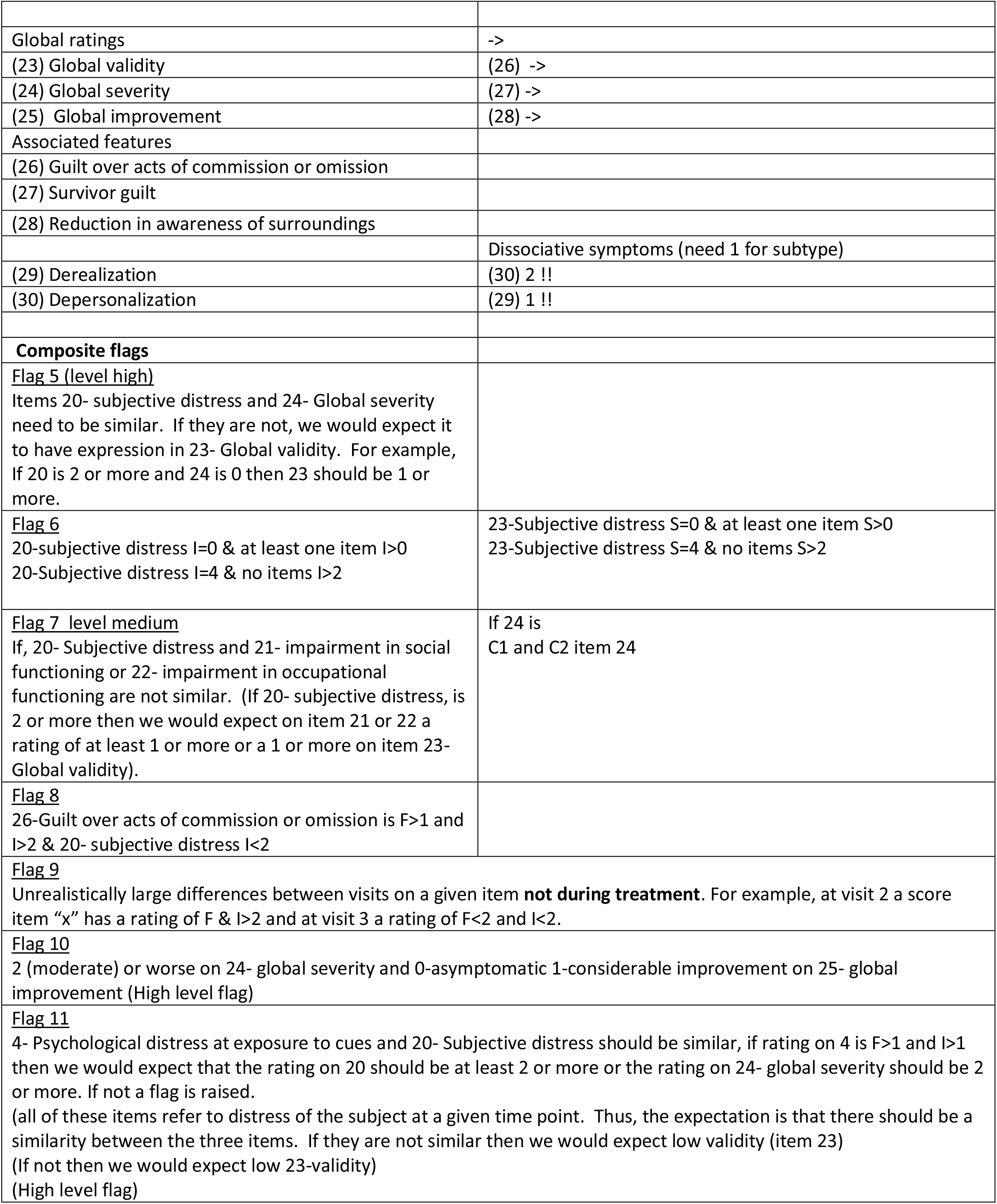
CAPS IV and CAPS V Inconsistency FLAGS.

## Conclusions

Scoring consistency flags for the CAPS may be useful in the quest to improve reliability and validity of clinical trials. Modified flags are currently being developed to cover the CAPS-V. Further testing of flags using clinical trial data is planned to examine their potential impact on signal detection.

## Data Availability

All data produced in the present work are contained in the manuscript.

## Acknowledgments

The research leading to these results has received support from the Elie Wiesel Chair at Bar Ilan University.

Previous versions of this material have been presented at ISCTM 2021 Autumn Conference

Jonathan Rabinowitz has received research grant(s) support and/or travel support and/or speaker fees and/or consultant fees from Janssen (J&J), Eli Lilly, Pfizer, Lundbeck, Roche, Abraham Pharmaceuticals, Pierre Fabre, Intra-cellular Therapies, Minerva, Takeda and Amgen. Alon A. Rabinowitz and Sara Freedman have no disclosures.

## References

Rabinowitz J, Schooler NR, Anderson A, et al. Consistency checks to improve measurement with the Positive and Negative Syndrome Scale (PANSS). Schizophr Res. 2017;190:74–76.

Rabinowitz, J., Schooler, NR, Brown, B. et al., Consistency checks to improve measurement with the Montgomery-Asberg Depression Rating Scale (MADRS), Journal of Affective Disorders 2019; 256: 143–147

Rabinowitz, J., Opler, M., Rabinowitz, A. A., Negash, S., Anderson, A., Fu, D. J., Williamson, D., Kott, A., Davis, L. L., & Schooler, N. R. (2021). Consistency checks to improve measurement with the Personal and Social Performance Scale (PSP). Schizophrenia research, 228, 529–533. 10.1016/j.schres.2020.11.040

Rabinowitz, J., Williams, J. B. W., Anderson, A., Fu, D. J., Hefting, N., Kadriu, B., Kott, A., Mahableshwarkar, A., Sedway, J., Williamson, D., Yavorsky, C., & Schooler, N. R. (2022). Consistency checks to improve measurement with the Hamilton Rating Scale for Depression (HAM-D). Journal of affective disorders, 302, 273–279. 10.1016/j.jad.2022.01.105

Rabinowitz, J., Young, R. C., Yavorsky, C., Williams, J. B. W., Sedway, J., Marino, P., Matteo, C., Mahableshwarkar, A., Kott, A., Hefting, N., Engler, J., & Brady, C. (2024). Consistency checks to improve measurement with the Young Mania Rating Scale (YMRS). Journal of affective disorders, 345, 24–31. 10.1016/j.jad.2023.10.098

Rabinowitz, J. & Rabinowitz, A. (2021). Outlier-response pattern checks to improve measurement with the Positive and Negative Syndrome Scale (PANSS). Psychiatry Research; Sep;303

Rabinowitz, J., & Rabinowitz, A. A. (2022). Outlier-response pattern checks to improve measurement with the Montgomery-Asberg depression rating scale (MADRS). Journal of affective disorders, 299, 444–448. 10.1016/j.jad.2021.12.076

